# Sex differences in the mortality rate for coronavirus disease 2019 compared to other causes of death

**DOI:** 10.1101/2021.02.23.21252314

**Authors:** P. Geldsetzer, T. Mukama, N. Jawad, T. Riffe, A. Rogers, N. Sudharsanan

## Abstract

Men are more likely than women to die due to coronavirus disease 2019 (COVID-19). This paper sets out to examine whether the magnitude of the sex differences in the COVID-19 mortality rate are unusual when compared to other common causes of death. In doing so, we aim to provide evidence as to whether the causal pathways for the sex differences in the mortality rate of COVID-19 likely differ from those for other causes of death. We found that sex differences in the age-standardized COVID-19 mortality rate were substantially larger than for the age-standardized all-cause mortality rate and most other common causes of death. These differences were especially large in the oldest age groups.

**One Sentence Summary:** The sex difference in the mortality rate of coronavirus disease 2019 is substantially larger than for other common causes of death.

## Introduction

Males have a higher risk of death from coronavirus disease 2019 (COVID-19) than females ^1–6^. This difference has been observed for both the case fatality rate (CFR; i.e., deaths among those diagnosed with a SARS-CoV-2 infection) and infection fatality rate (IFR; i.e., deaths among all those who were infected with SARS-CoV-2) ^1^. This higher risk of death from COVID-19 in males has been highlighted both in the academic literature and the media ^7–9^.

Understanding why these disparities by sex exist has become an active area of research. However, given that the risk of death from COVID-19 is strongly related to one’s expected remaining life expectancy ^6^, it is unclear whether the observed sex differences in the COVID-19 fatality rate are simply a reflection of men’s shorter life expectancy ^10^, which is at least in part due to their poorer health status at any given age. This study aimed to determine if sex differences in COVID-19 mortality are larger when compared to the all-cause mortality rate, mortality rates for other common causes of death, and – given SARS-CoV-2’s common respiratory manifestations – other respiratory causes of death, including respiratory infections. This information is crucial, as it begins to elucidate whether the higher COVID-19 mortality risk among males reflects the survival advantage among females compared to males, and is, thus, likely a result of the biological, behavioral, and social pathways that cause this survival advantage as opposed to causal pathways that are specific to COVID-19. Understanding these causal pathways could help in the development of therapeutics and preventive measures for COVID-19 and future coronavirus disease outbreaks.

## Results

We extracted the latest available country-level data on COVID-19 deaths from the COVerAGE-DB for countries for which age- and sex-disaggregated data were available (as of 09 February 2021) ^11^. We then obtained age- and sex-disaggregated data on all-cause mortality and population size for each of these countries from the Human Mortality Database (HMD) ^12^ and, for countries not included in the HMD, from the United Nation’s World Population Prospects (WPP) ^10^. The latest available mortality data for specific causes of death were drawn from the World Health Organization’s (WHO) mortality database ^13^. We then calculated the ratios for the sex difference in the COVID-19 mortality (i.e., the ratio of the number of COVID-19 deaths in men divided by the male mid-year population to the number of COVID-19 deaths in women divided by the female mid-year population) and compared these ratios to those for i) all-cause mortality, ii) each of the six most common causes of death groups globally for adults (excluding injuries) ^14^, and iii) each major respiratory cause of death.

Age- and sex-disaggregated data on COVID-19 deaths were available for 59 countries (Table 1).

**Table 1.**
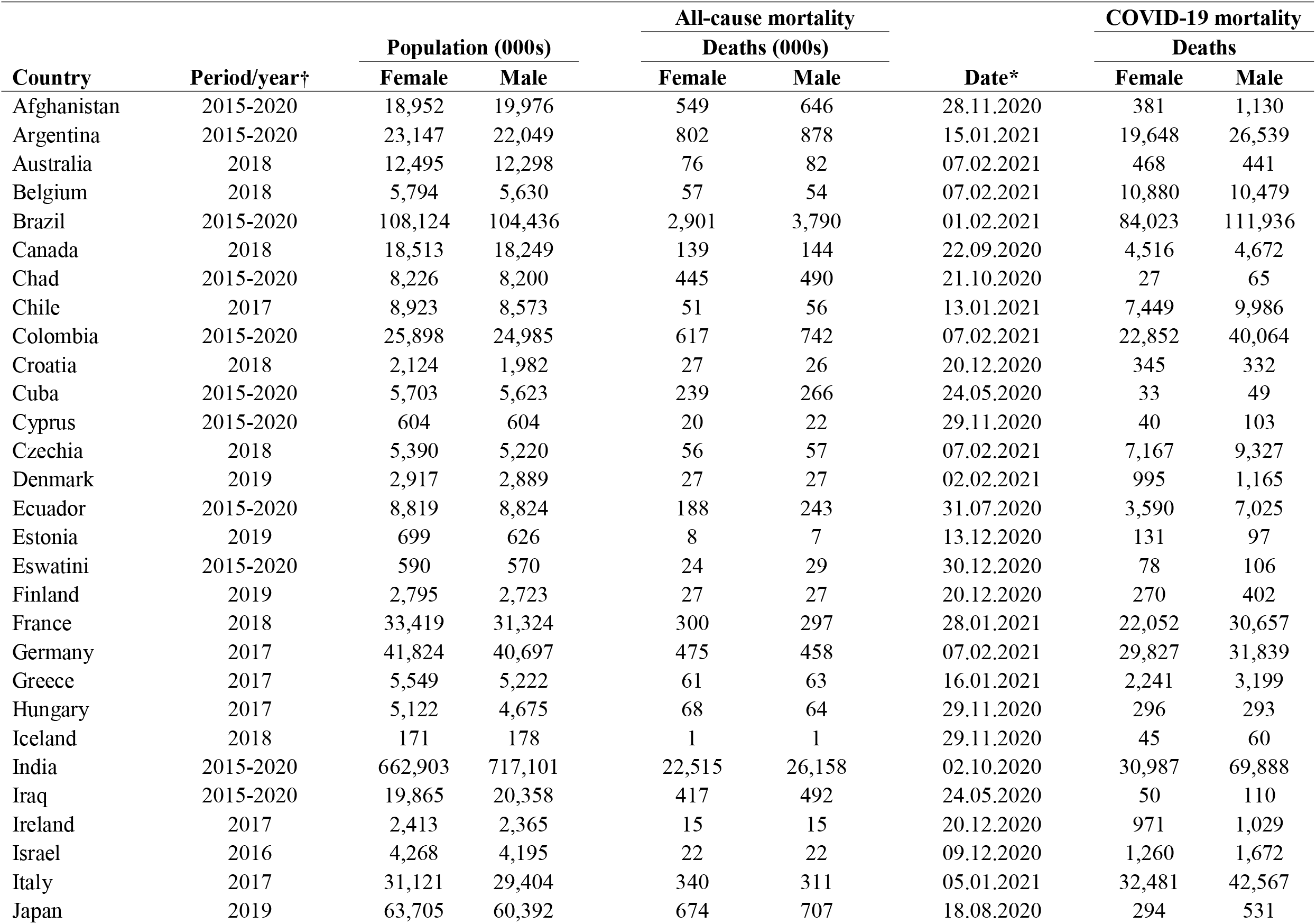
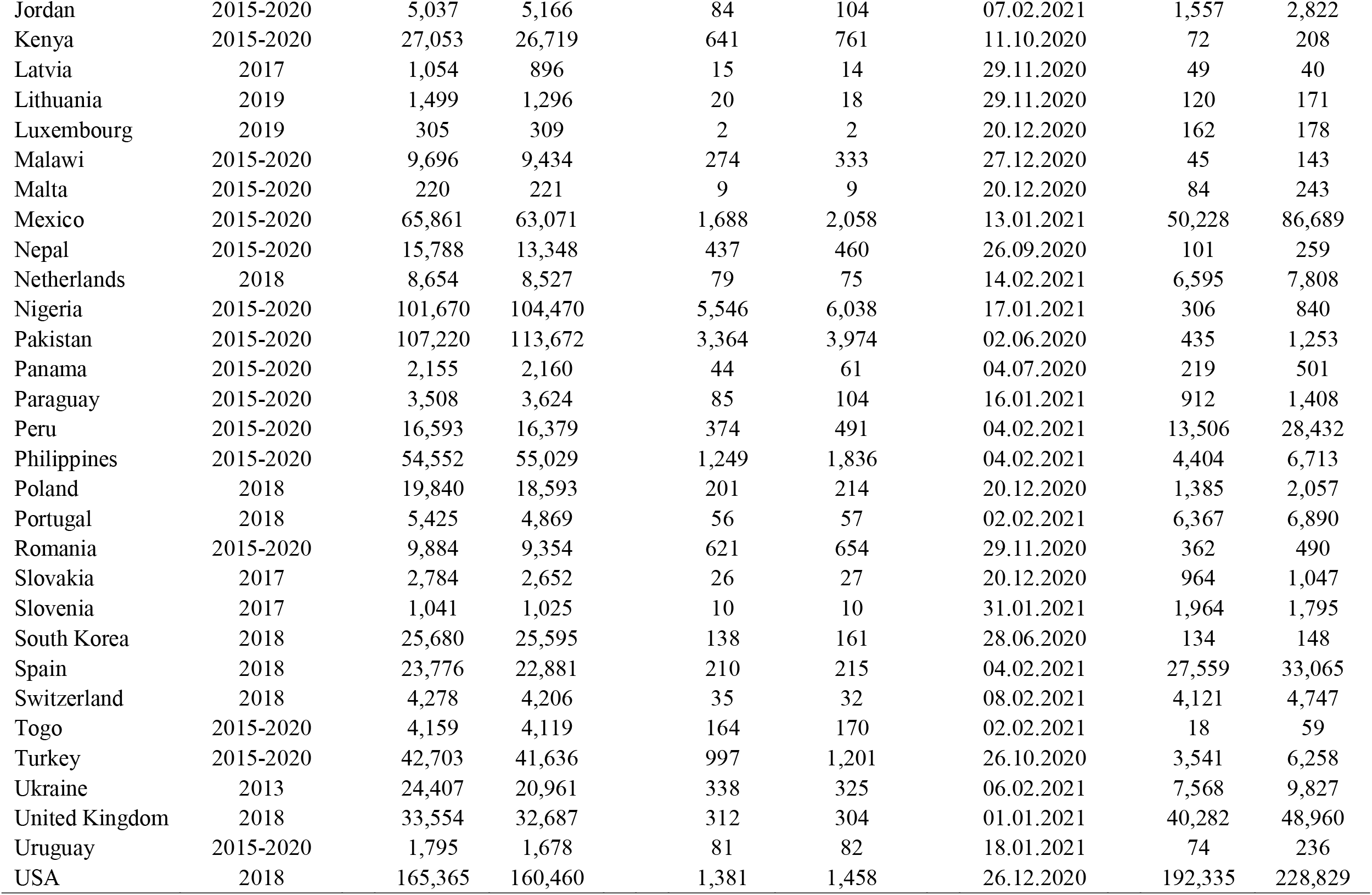
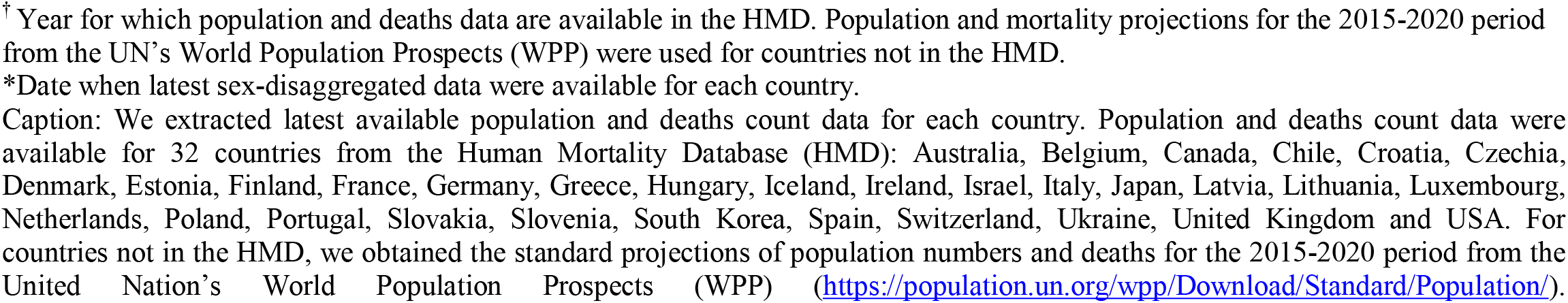
Population, all-cause deaths, and COVID-19 cases and deaths by sex and country.

| Country | Period/year† | Population (000s) |  | All-cause mortality |  | Date* | COVID-19 mortality |  |
| --- | --- | --- | --- | --- | --- | --- | --- | --- |
|  |  | Female | Male | Deaths (000s) |  |  | Deaths |  |
|  |  |  |  | Female | Male |  | Female | Male |
| Afghanistan | 2015-2020 | 18,952 | 19,976 | 549 | 646 | 28.11.2020 | 381 | 1,130 |
| Argentina | 2015-2020 | 23,147 | 22,049 | 802 | 878 | 15.01.2021 | 19,648 | 26,539 |
| Australia | 2018 | 12,495 | 12,298 | 76 | 82 | 07.02.2021 | 468 | 441 |
| Belgium | 2018 | 5,794 | 5,630 | 57 | 54 | 07.02.2021 | 10,880 | 10,479 |
| Brazil | 2015-2020 | 108,124 | 104,436 | 2,901 | 3,790 | 01.02.2021 | 84,023 | 111,936 |
| Canada | 2018 | 18,513 | 18,249 | 139 | 144 | 22.09.2020 | 4,516 | 4,672 |
| Chad | 2015-2020 | 8,226 | 8,200 | 445 | 490 | 21.10.2020 | 27 | 65 |
| Chile | 2017 | 8,923 | 8,573 | 51 | 56 | 13.01.2021 | 7,449 | 9,986 |
| Colombia | 2015-2020 | 25,898 | 24,985 | 617 | 742 | 07.02.2021 | 22,852 | 40,064 |
| Croatia | 2018 | 2,124 | 1,982 | 27 | 26 | 20.12.2020 | 345 | 332 |
| Cuba | 2015-2020 | 5,703 | 5,623 | 239 | 266 | 24.05.2020 | 33 | 49 |
| Cyprus | 2015-2020 | 604 | 604 | 20 | 22 | 29.11.2020 | 40 | 103 |
| Czechia | 2018 | 5,390 | 5,220 | 56 | 57 | 07.02.2021 | 7,167 | 9,327 |
| Denmark | 2019 | 2,917 | 2,889 | 27 | 27 | 02.02.2021 | 995 | 1,165 |
| Ecuador | 2015-2020 | 8,819 | 8,824 | 188 | 243 | 31.07.2020 | 3,590 | 7,025 |
| Estonia | 2019 | 699 | 626 | 8 | 7 | 13.12.2020 | 131 | 97 |
| Eswatini | 2015-2020 | 590 | 570 | 24 | 29 | 30.12.2020 | 78 | 106 |
| Finland | 2019 | 2,795 | 2,723 | 27 | 27 | 20.12.2020 | 270 | 402 |
| France | 2018 | 33,419 | 31,324 | 300 | 297 | 28.01.2021 | 22,052 | 30,657 |
| Germany | 2017 | 41,824 | 40,697 | 475 | 458 | 07.02.2021 | 29,827 | 31,839 |
| Greece | 2017 | 5,549 | 5,222 | 61 | 63 | 16.01.2021 | 2,241 | 3,199 |
| Hungary | 2017 | 5,122 | 4,675 | 68 | 64 | 29.11.2020 | 296 | 293 |
| Iceland | 2018 | 171 | 178 | 1 | 1 | 29.11.2020 | 45 | 60 |
| India | 2015-2020 | 662,903 | 717,101 | 22,515 | 26,158 | 02.10.2020 | 30,987 | 69,888 |
| Iraq | 2015-2020 | 19,865 | 20,358 | 417 | 492 | 24.05.2020 | 50 | 110 |
| Ireland | 2017 | 2,413 | 2,365 | 15 | 15 | 20.12.2020 | 971 | 1,029 |
| Israel | 2016 | 4,268 | 4,195 | 22 | 22 | 09.12.2020 | 1,260 | 1,672 |
| Italy | 2017 | 31,121 | 29,404 | 340 | 311 | 05.01.2021 | 32,481 | 42,567 |
| Japan | 2019 | 63,705 | 60,392 | 674 | 707 | 18.08.2020 | 294 | 531 |
| Jordan | 2015-2020 | 5,037 | 5,166 | 84 | 104 | 07.02.2021 | 1,557 | 2,822 |
| Kenya | 2015-2020 | 27,053 | 26,719 | 641 | 761 | 11.10.2020 | 72 | 208 |
| Latvia | 2017 | 1,054 | 896 | 15 | 14 | 29.11.2020 | 49 | 40 |
| Lithuania | 2019 | 1,499 | 1,296 | 20 | 18 | 29.11.2020 | 120 | 171 |
| Luxembourg | 2019 | 305 | 309 | 2 | 2 | 20.12.2020 | 162 | 178 |
| Malawi | 2015-2020 | 9,696 | 9,434 | 274 | 333 | 27.12.2020 | 45 | 143 |
| Malta | 2015-2020 | 220 | 221 | 9 | 9 | 20.12.2020 | 84 | 243 |
| Mexico | 2015-2020 | 65,861 | 63,071 | 1,688 | 2,058 | 13.01.2021 | 50,228 | 86,689 |
| Nepal | 2015-2020 | 15,788 | 13,348 | 437 | 460 | 26.09.2020 | 101 | 259 |
| Netherlands | 2018 | 8,654 | 8,527 | 79 | 75 | 14.02.2021 | 6,595 | 7,808 |
| Nigeria | 2015-2020 | 101,670 | 104,470 | 5,546 | 6,038 | 17.01.2021 | 306 | 840 |
| Pakistan | 2015-2020 | 107,220 | 113,672 | 3,364 | 3,974 | 02.06.2020 | 435 | 1,253 |
| Panama | 2015-2020 | 2,155 | 2,160 | 44 | 61 | 04.07.2020 | 219 | 501 |
| Paraguay | 2015-2020 | 3,508 | 3,624 | 85 | 104 | 16.01.2021 | 912 | 1,408 |
| Peru | 2015-2020 | 16,593 | 16,379 | 374 | 491 | 04.02.2021 | 13,506 | 28,432 |
| Philippines | 2015-2020 | 54,552 | 55,029 | 1,249 | 1,836 | 04.02.2021 | 4,404 | 6,713 |
| Poland | 2018 | 19,840 | 18,593 | 201 | 214 | 20.12.2020 | 1,385 | 2,057 |
| Portugal | 2018 | 5,425 | 4,869 | 56 | 57 | 02.02.2021 | 6,367 | 6,890 |
| Romania | 2015-2020 | 9,884 | 9,354 | 621 | 654 | 29.11.2020 | 362 | 490 |
| Slovakia | 2017 | 2,784 | 2,652 | 26 | 27 | 20.12.2020 | 964 | 1,047 |
| Slovenia | 2017 | 1,041 | 1,025 | 10 | 10 | 31.01.2021 | 1,964 | 1,795 |
| South Korea | 2018 | 25,680 | 25,595 | 138 | 161 | 28.06.2020 | 134 | 148 |
| Spain | 2018 | 23,776 | 22,881 | 210 | 215 | 04.02.2021 | 27,559 | 33,065 |
| Switzerland | 2018 | 4,278 | 4,206 | 35 | 32 | 08.02.2021 | 4,121 | 4,747 |
| Togo | 2015-2020 | 4,159 | 4,119 | 164 | 170 | 02.02.2021 | 18 | 59 |
| Turkey | 2015-2020 | 42,703 | 41,636 | 997 | 1,201 | 26.10.2020 | 3,541 | 6,258 |
| Ukraine | 2013 | 24,407 | 20,961 | 338 | 325 | 06.02.2021 | 7,568 | 9,827 |
| United Kingdom | 2018 | 33,554 | 32,687 | 312 | 304 | 01.01.2021 | 40,282 | 48,960 |
| Uruguay | 2015-2020 | 1,795 | 1,678 | 81 | 82 | 18.01.2021 | 74 | 236 |
| USA | 2018 | 165,365 | 160,460 | 1,381 | 1,458 | 26.12.2020 | 192,335 | 228,829 |

<sup>†</sup> Year for which population and deaths data are available in the HMD. Population and mortality projections for the 2015-2020 period from the UN's World Population Prospects (WPP) were used for countries not in the HMD.
\*Date when latest sex-disaggregated data were available for each country.
5 Caption: We extracted latest available population and deaths count data for each country. Population and deaths count data were available for 32 countries from the Human Mortality Database (HMD): Australia, Belgium, Canada, Chile, Croatia, Czechia, Denmark, Estonia, Finland, France, Germany, Greece, Hungary, Iceland, Ireland, Israel, Italy, Japan, Latvia, Lithuania, Luxembourg, Netherlands, Poland, Portugal, Slovakia, Slovenia, South Korea, Spain, Switzerland, Ukraine, United Kingdom and USA. For countries not in the HMD, we obtained the standard projections of population numbers and deaths for the 2015-2020 period from the United Nation's World Population Prospects (WPP) (https://population.un.org/wpp/Download/Standard/Population/).

From this data, we calculated the age-standardized male-to-female rate ratios of mortality from COVID-19 and all-causes. The age standardization, conducted separately for each country, was carried out to ensure that differences in the male-to-female mortality rate ratio between COVID-19 and all causes were not confounded by sex differences in the age distribution. Point estimates greater than one in Figure 1 indicate that men had a higher rate of death than women. We found that in most countries, the male disadvantage for COVID-19 mortality was substantially larger than their all-cause mortality disadvantage (Figure 1 and Figure S1).

**Figure 1.**
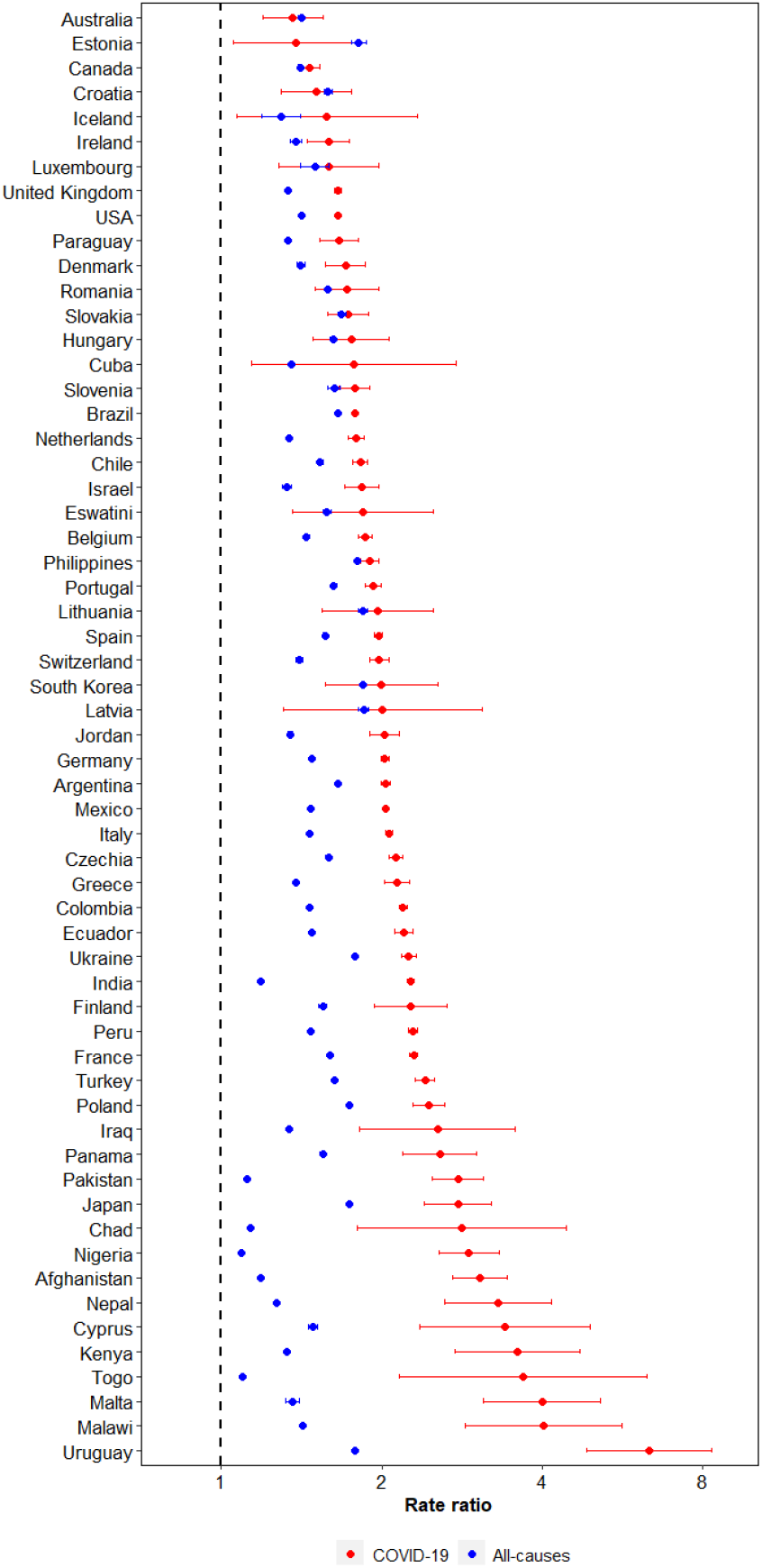
Male-to-female rate ratios of mortality from COVID-19 and all causes. Caption: Rate ratios for the sex differences in COVID-19 and all-cause mortality were calculated for each country by dividing the age-standardized mortality rate in males by the age-standardized mortality rate in females.

Next, we investigated whether the degree of the male disadvantage in the COVID-19 compared to the all-cause mortality varied by age group. To do so, we divided – separately by ten-year age group – the age-standardized male-to-female rate ratio for the COVID-19 mortality by the age-standardized male-to-female rate ratio for all-cause mortality. Point estimates greater than one in Figure 2, thus, indicate that the sex differences in the mortality rate for COVID-19 were greater than expected based on the sex differences in the all-cause mortality rate. We found that among the older age groups, especially the group aged 80 years and older, the higher rate of death for men than woman exceeded (in relative terms) that for all-cause mortality for most countries. Among younger age groups, especially those aged less than 50 years, the direction and magnitude of these differences varied greatly by country. These patterns are similar when adjusting these estimates for remaining life expectancy (Figure S2).

**Figure 2.**
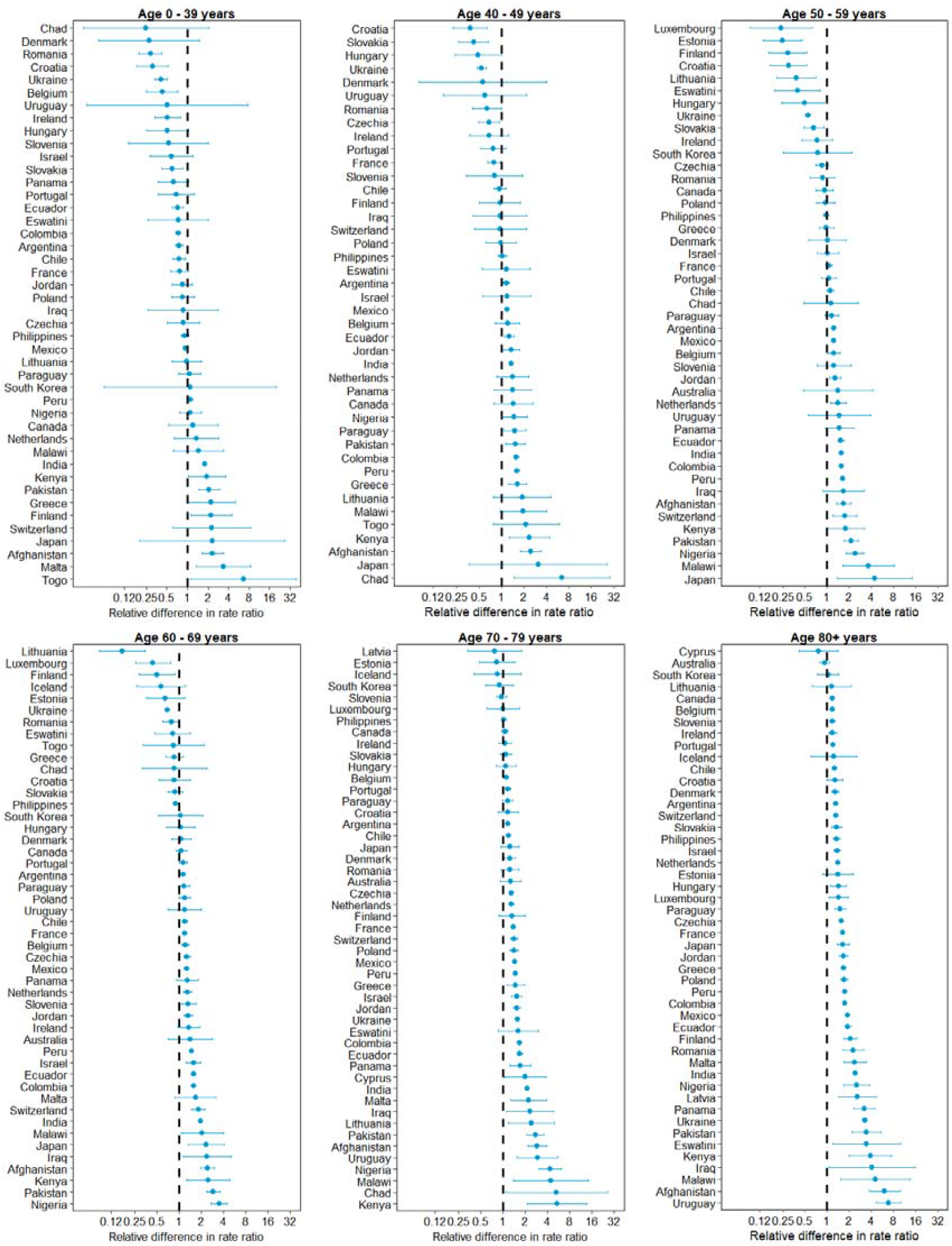
Relative difference in the male-to-female rate ratios of COVID-19-specific and all-cause mortality, by age group. Caption: The relative difference in the rate ratio was calculated by dividing (separately among each age group shown) the male-to-female rate ratio for the COVID-19-specific mortality rate by the male-to-female rate ratio for the all-cause mortality rate.

Using the same metric as for Figure 2, we then compared the relative magnitude of sex differences in the mortality for COVID-19 to that for other major causes of mortality (circulatory diseases, cancer, chronic respiratory diseases, respiratory infections and tuberculosis, diabetes, and neurologic disorders). We found that in most countries the relative sex differences for COVID-19 were larger than for each of the other common causes of death (Figure 3 and Figure S3). However, this was not true for chronic respiratory conditions for which countries were spread approximately equally across the vertical dashed line drawn at one (i.e., the number indicating that the relative sex difference for the COVID-19 mortality was the same as for chronic respiratory diseases). Implementing the same analysis as in Figure 3 for each common respiratory cause of death (the ICD-10 codes used for categorization are shown in Table S2) revealed that the similar male disadvantage in mortality for chronic respiratory diseases as for COVID-19 is largely driven by a high male disadvantage in mortality from bronchitis and emphysema (Figure S4) and, thus, likely the higher prevalence of smoking (especially in the past) among men than women ^15,16^.

**Figure 3.**
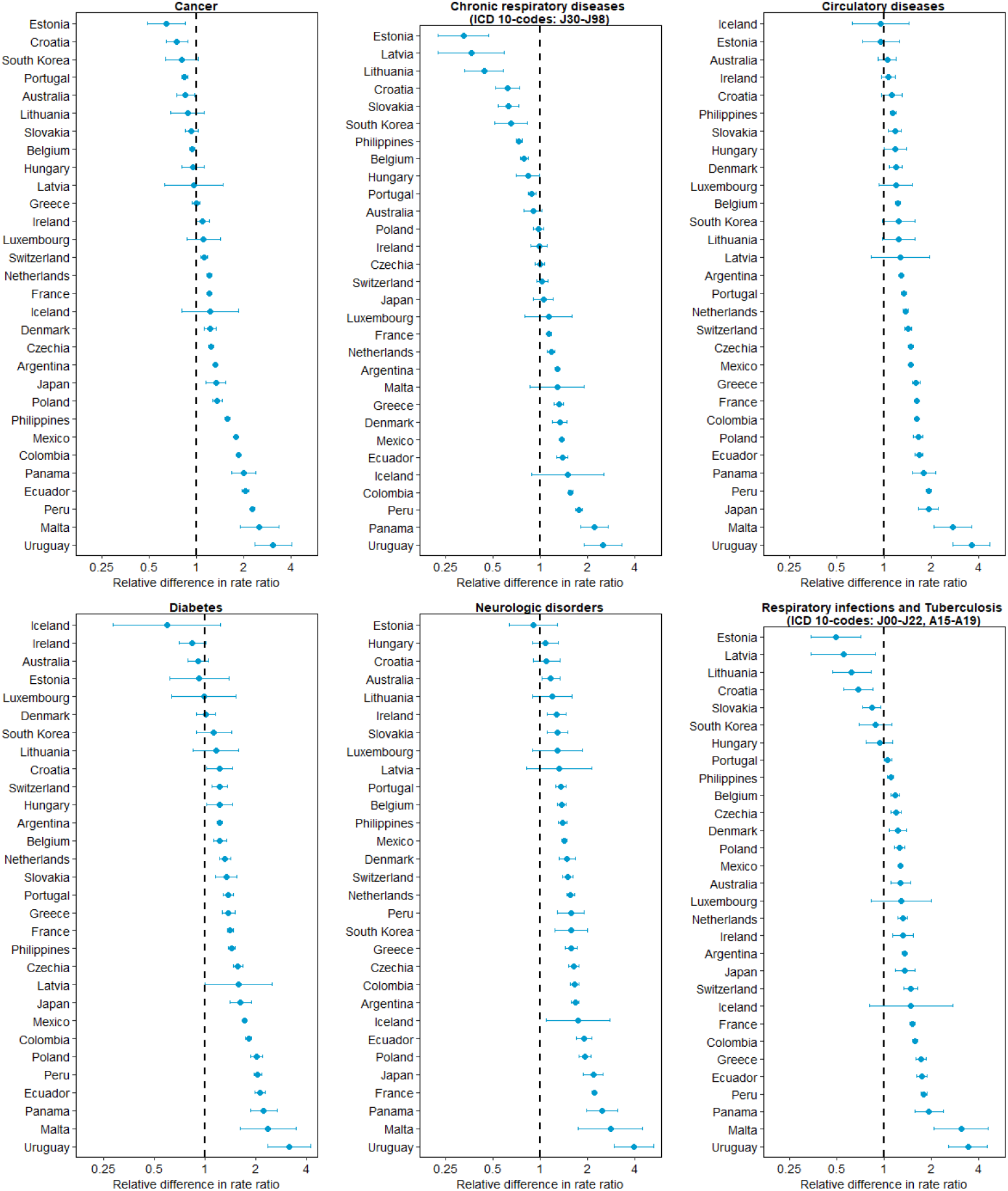
Relative difference in the male-to-female rate ratios of COVID-19-specific mortality and six major causes of mortality.

## Discussion

The degree to which men are more likely to die from COVID-19 than women is substantially larger than would be expected merely based on the fact that men are generally more likely to die at any given age than women. Thus, the probability of succumbing to a SARS-CoV-2 infection does not appear to be fully explained by the remaining life expectancy of the person who was infected. This observation suggests that the causal pathways that link male sex to a shorter life expectancy may not fully explain the unusually high male disadvantage in COVID-19 mortality. Our findings, therefore, lend support to hypotheses that posit that the causal pathways that link male sex to a higher mortality from a SARS-CoV-2 infection are specific to SARS-CoV-2 rather than shared with the pathways responsible for the shorter life expectancy among men than women or the causal pathways for sex differences for other common causes of death.

This study has several limitations. First and foremost, this study can only provide suggestive (as opposed to conclusive) evidence as to whether or not the causal pathways underlying the male disadvantage for COVID-19 mortality are shared with those underlying the all-cause mortality disadvantage for men. Second, our mortality rate calculations for COVID-19 use the total population (by sex) as the denominator. Thus, the assumption underlying the validity of our calculation is that there are no substantial differences in the probability of being infected with SARS-CoV-2 between males and females. To date, evidence from seroprevalence studies suggests that this assumption is reasonable ^17,18^. An alternative approach is to use the number of identified cases of SARS-CoV-2 infections as the denominator (i.e., calculating the case fatality rate). This approach, however, assumes that the degree of underdetection of SARS-CoV-2 infections is the same among men as among women. This assumption would, for example, be violated if males are more likely to develop symptoms from a SARS-CoV-2 infection than females and are, therefore, more likely to seek out a COVID-19 test, or if men have better access to testing than women. Although both choices for the denominator (total population or number of cases) rely on untestable assumptions, our analyses in which we use the number of cases instead of the total population as denominator found that the choice of denominator does not substantially change our conclusions.

Studies have hypothesized that the sex differences in COVID-19 mortality exist due to behavioral and social risk factors (e.g., higher incidence of smoking and drinking among men than women) that place men at a greater risk of mortality from health complications associated with COVID-19 ^19–24^. Other studies have cited a higher rate of comorbidities, such as diabetes and heart disease, as the reason for the higher COVID-19 fatality rate among men ^4,25–28^. Finally, some studies suggest biological factors that may explain these disparities. One potential factor is a higher expression among men than women of the angiotensin-converting enzyme 2 receptor, which is used by SARS-CoV-2 to enter the host cell ^3,29,30^. Other possible biological factors relate to immunological differences between males and females ^31–33^. Ultimately a combination of biological, behavioral, and social pathways may be responsible for the high male disadvantage in COVID-19 mortality. Elucidating these causal chains is an important research area given that it may assist in the development of therapeutics and preventive measures for COVID-19 and future outbreaks of coronavirus diseases.

## Data Availability

All the data used in the study are publicly available. COVID-19 mortality data are available from the COVerAGE-DB. Data on population size by age and sex are available from the HMD and the United Nations WPP.

https://osf.io/mpwjq/

https://www.mortality.org/

https://www.who.int/healthinfo/mortality_data/en/

## Author contributions

Pascal Geldsetzer: conceptualization, methodology, writing – original draft preparation; Trasias Mukama: methodology, data analysis, writing – reviewing and editing; Nadine Jawad: methodology, writing – original draft preparation; Tim Riffe: conceptualization, methodology, writing – reviewing and editing; Angela Rogers: methodology, writing – reviewing and editing; Nikkil Sudharsanan: conceptualization, methodology, writing – reviewing and editing;

## Data and materials availability

All the data used in the study are publicly available. COVID-19 mortality data are available from the COVerAGE-DB (https://osf.io/mpwjq/). Data on population size by age and sex are available from the HMD (www.mortality.org) and the United Nation’s WPP (https://www.who.int/healthinfo/mortality_data/en/).

