## Supplemental Figures for "Sex differences in the mortality rate for coronavirus disease 2019 compared to other causes of death"

**This PDF file includes:**

Materials and Methods

Supplementary Text

Figs. S1 to S4

Table S1

Materials and Methods

**Data sources**

***COVID-19 mortality data***

We extracted the latest available country-level data on COVID-19 deaths from the COVerAGE-DB for countries for which age- and sex-disaggregated data were available (as of 09 February 2021) and which had at least 50 COVID-19 deaths as of 09 February 2021 *(10).* Age- and sex-disaggregated data were available for 59 countries: Afghanistan, Argentina, Australia, Belgium, Brazil, Canada, Chad, Chile, Colombia, Croatia, Cuba, Cyprus, Czechia, Denmark, Ecuador, Estonia, Eswatini, Finland, France, Germany, Greece, Hungary, Iceland, India, Iraq, Ireland, Israel, Italy, Japan, Jordan, Kenya, Latvia, Lithuania, Luxembourg, Malawi, Malta, Mexico, Nepal, Netherlands, Nigeria, Pakistan, Panama, Paraguay, Peru, Philippines, Poland, Portugal, Romania, Slovakia, Slovenia, South Korea, Spain, Switzerland, Togo, Turkey, Ukraine, United Kingdom, Uruguay, and USA. The disaggregation by age in the COVerAGE-DB was by ten-year age group.

***General mortality data***

For the 59 countries for which sex- and age-disaggregated data was available, we obtained age- and sex-disaggregated data on all-cause mortality and total population size from the Human Mortality Database (HMD) ^12^We extracted the latest available population and deaths count data for each country (as of 09 February 2021). Population and deaths count data were available for 32 countries from the HMD: Australia, Belgium, Canada, Chile, Croatia, Czechia, Denmark, Estonia, Finland, France, Germany, Greece, Hungary, Iceland, Ireland, Israel, Italy, Japan, Latvia, Lithuania, Luxembourg, Netherlands, Poland, Portugal, Slovakia, Slovenia, South Korea, Spain, Switzerland, Ukraine, United Kingdom and USA.

For countries not in the HMD, we obtained the standard projections of populations and deaths data for the 2015-2020 period from the United Nation’s World Population Prospects (WPP) *(12)* . Data on population size and deaths were available for one-year age groups in the HMD and for 5-year age categories in the WPP. We categorised data from both the HMD and WPP into ten-year age groups, and combined data for age groups older than 80 into one category (80+).

***Cause-specific mortality data***

We also obtained the latest available mortality data for specific causes of deaths from the WHO mortality database *(13).* Causes of death are classified according to the 10^th^ revision of the International Classification of Diseases (ICD-10) (*16*). We obtained data for the top six causes of death groups globally (according to the WHO mortality database *(13)*), which were circulatory diseases, cancer, chronic respiratory diseases, respiratory infections and tuberculosis, diabetes, and neurologic disorders. To compare COVID-19 mortality differences by sex with differences observed for other respiratory causes of deaths, we further categorized respiratory causes into 6 groups: acute upper respiratory infections, influenza, pneumonia, other acute lower respiratory infections, other diseases of the upper respiratory tract, and chronic lower respiratory diseases. The ICD-10 codes used to define the 6 major causes of mortality and the respiratory causes of death are presented in Table S3.

**Analysis**

We calculated age-standardized mortality rates separately for each country for COVID-19-specific, cause-specific, and all-cause mortality. We first estimated COVID-19, cause-specific, and all-cause mortality rates for each age group by dividing the total number of deaths due to each cause (or all deaths for all-cause) by the mid-year population in that age group. To allow more robust between-sex rate ratios within countries, we then estimated sex-specific age-standardized mortality rates for each country using the overall age distribution of each country as the standard.

Rate ratios for the sex differences in COVID-19 mortality were calculated for each country by dividing the age-standardized COVID-19-specific mortality rate in men by the age-standardized COVID-19-specific mortality rate in women. Similarly, sex differences in all-cause mortality were examined using the rate ratios obtained by dividing the age-standardized all-cause mortality rate in men by the age-standardized all-cause mortality rate in women. We calculated excess mortality – the difference between the male to female mortality rate ratio from COVID-19 and that from all causes – by dividing the two ratios. We present the relative difference in these rate ratios of mortality by age group (0-39, 40-49, 50-59, 60-69, 70-79, and 80+ years), major causes of mortality (circulatory diseases, cancer, chronic respiratory diseases, respiratory infections and tuberculosis, diabetes, and neurologic disorders), and common respiratory causes of death.

Lastly, instead of using age standardization, we calculated remaining life expectancy-adjusted mortality rates for men and women. The rationale for this robustness check is that remaining life expectancy may be a better measure of biological age than calendar age. We obtained data on remaining life expectancy by age from the HMD and WPP. Specifically, for the latest year (in HMD) and period (in the WPP) for which data were available as of 09 February 2021, we obtained the remaining life expectancy at each single year of age separately for each country and sex. Again separately for each country and sex, we then categorized remaining life expectancy as follows: 0-4, 5-9, 10-14, 15-19, 20-24, 25-29, 30-34, 35-39, 40-44, 45-50, and 50+ years. Thus, it was possible for men and women of the same age in the same country to belong to different remaining life-expectancy groups. We then applied the same approach as described above for age standardization to calculate a weighted sum of the remaining life expectancy group-specific mortality rate. We refer to this weighted sum as the remaining life expectancy-adjusted mortality rates.

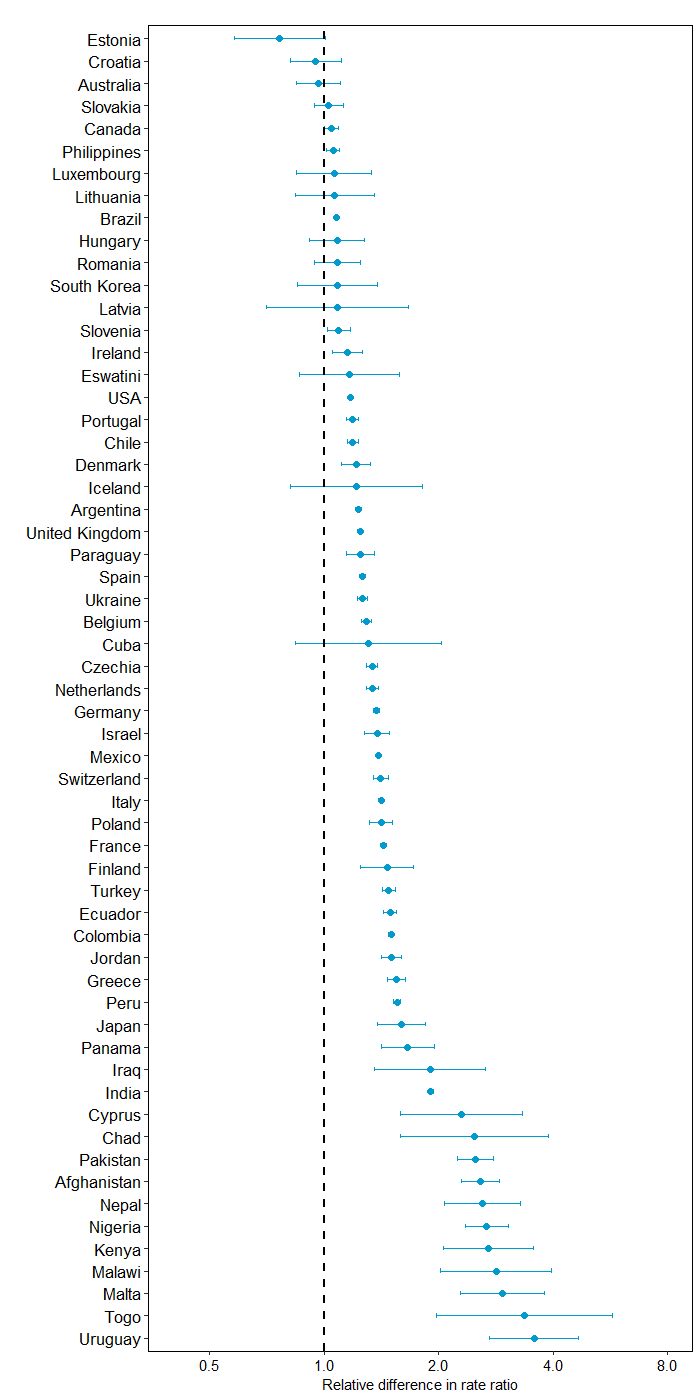

Fig. S1. Relative difference in the male-to-female rate ratios of COVID-19-specific and all-cause mortality

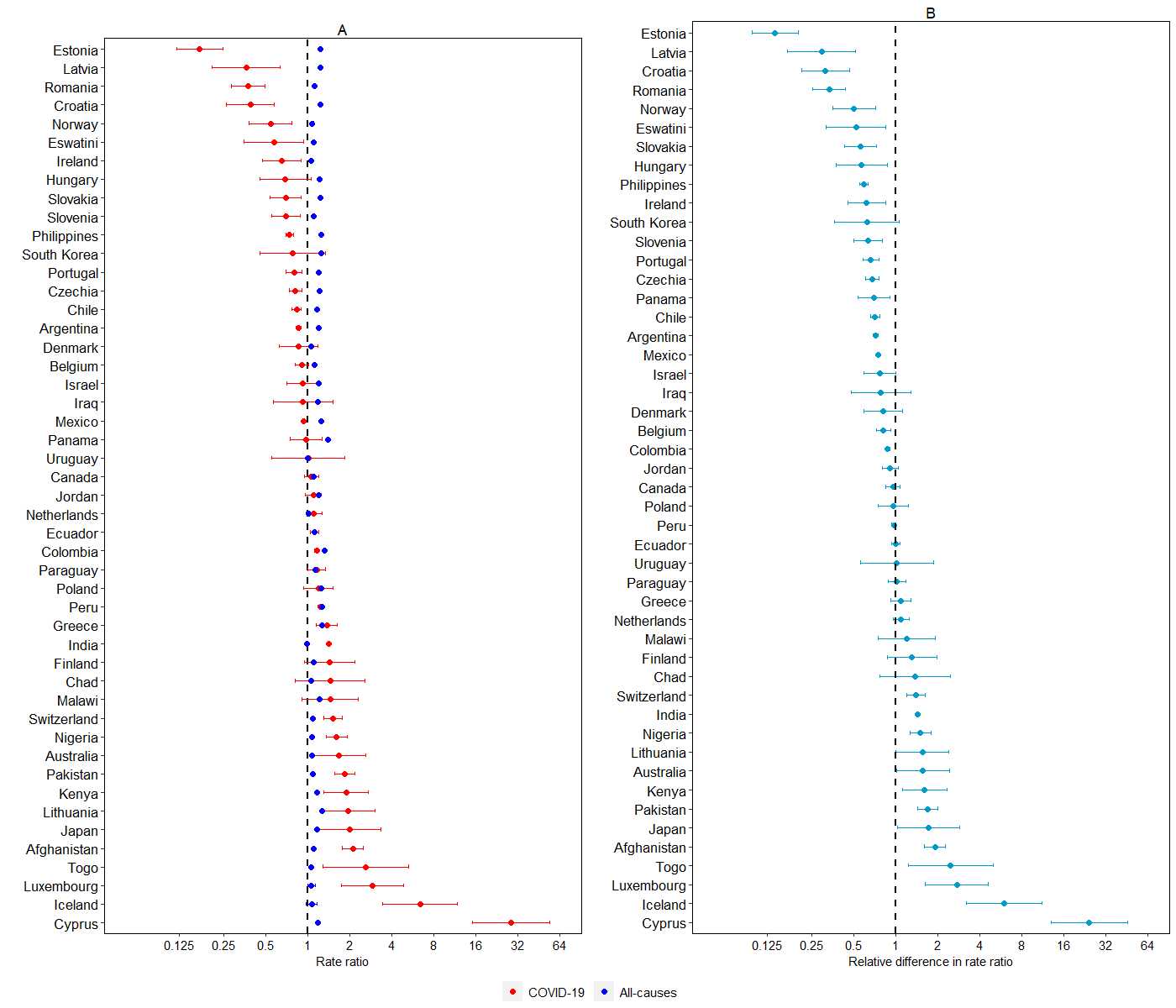

Fig. S2. Panel A: Male-to-female rate ratios of mortality from COVID-19 and all-cause deaths adjusted for remaining life expectancy. Panel B: Relative difference in the male-to-female rate ratios of COVID-19-specific and all-cause mortality adjusted for remaining life expectancy.

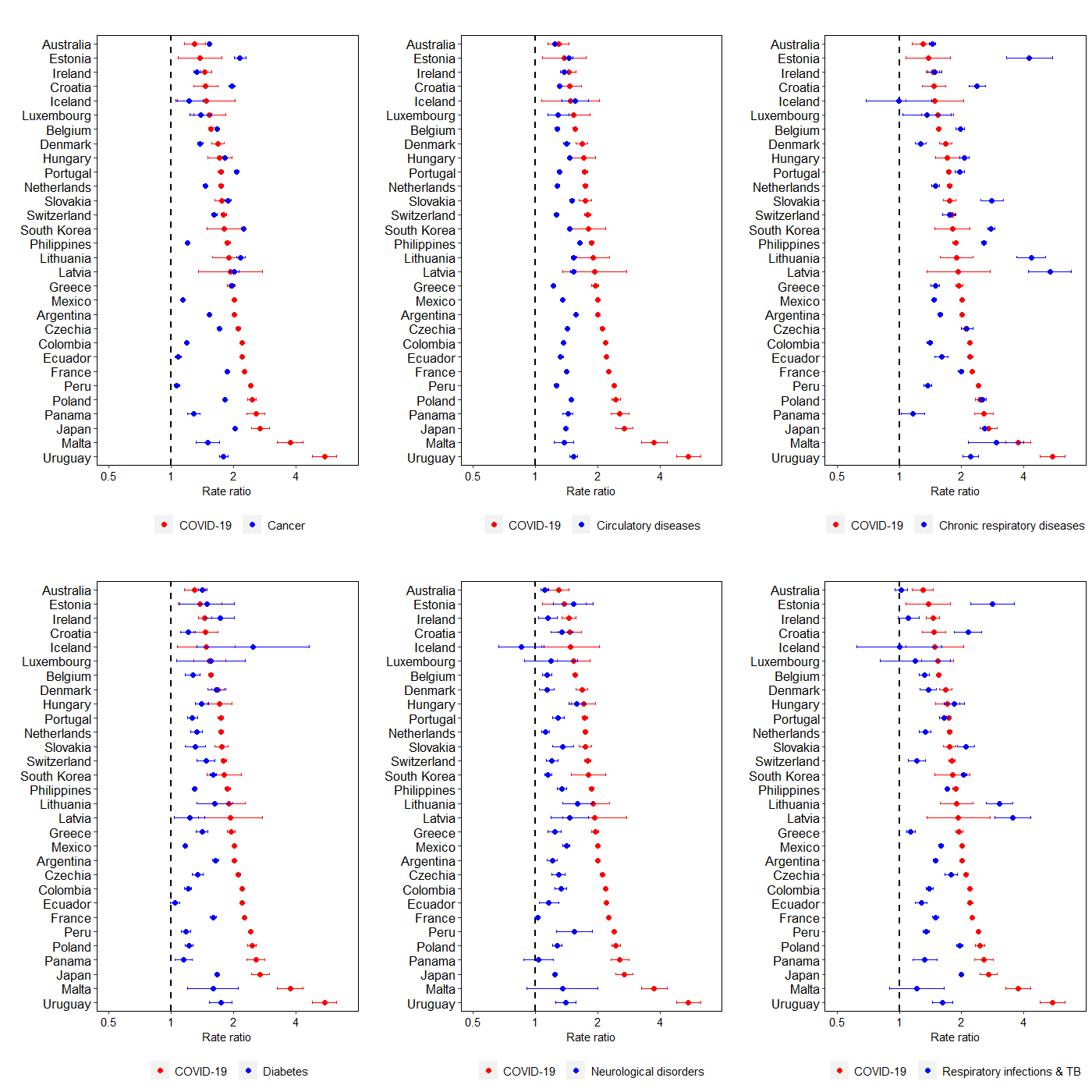

Fig. S3. Male-to-female rate ratios of mortality from COVID-19 and six major causes of mortality.

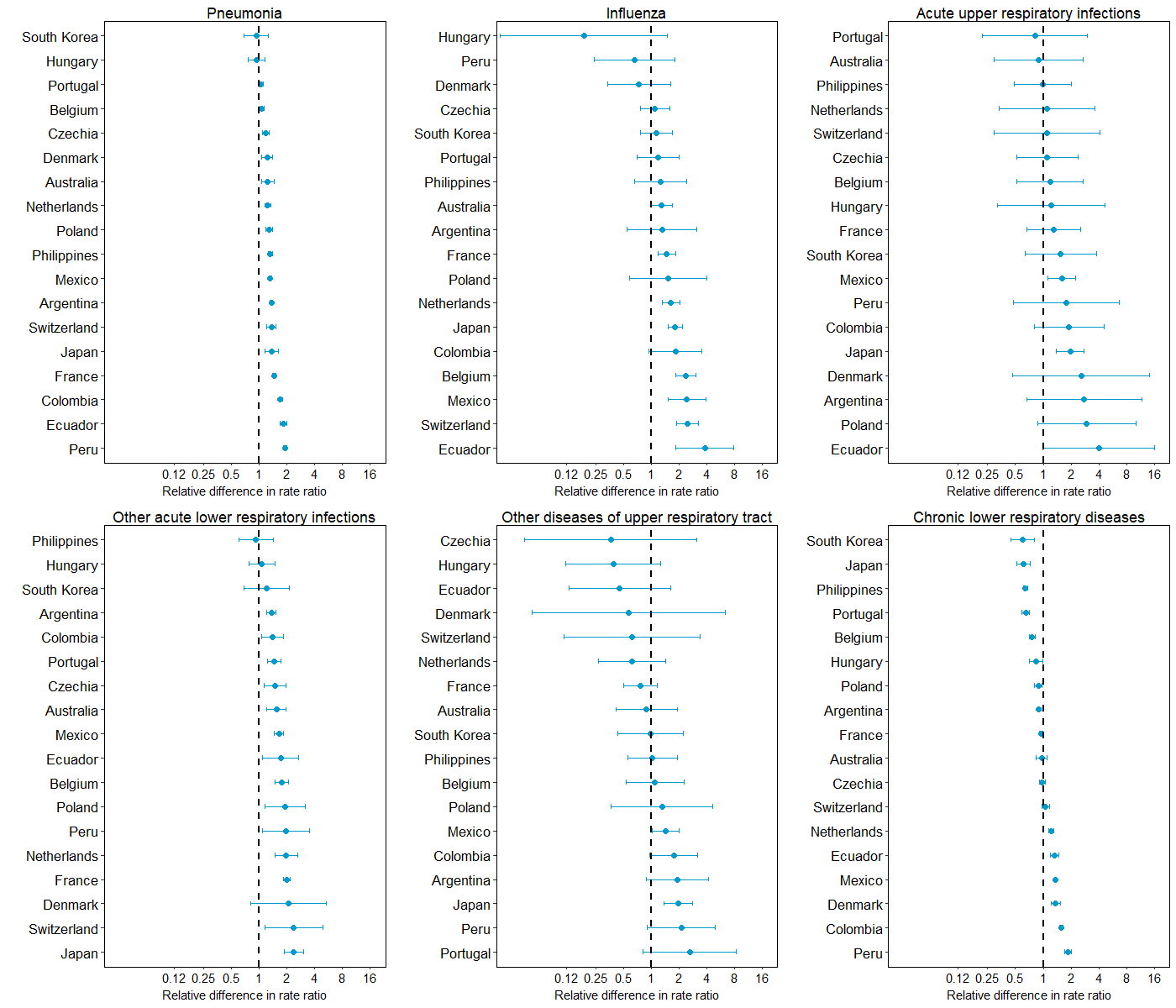

Fig. S4. Relative difference in the male-to-female rate ratios of mortality from COVID-19 and other respiratory diseases

Caption: The ICD-10 codes corresponding to each respiratory disease are shown in Table S2.

Table S1. Age- and sex-disaggregated data on six major causes of deaths

|  |  |  | **Mortality (number of deaths)** | | | | | | | | | | | | | | | | |
| --- | --- | --- | --- | --- | --- | --- | --- | --- | --- | --- | --- | --- | --- | --- | --- | --- | --- | --- | --- |
|  |  |  | **Cancer** | |  | **Chronic respiratory diseases** | |  | **Circulatory diseases** | |  | **Diabetes** | |  | **Neurologic conditions** | |  | **Respiratory infections and TB** | |
| **Country** | **Year** |  | **Female** | **Male** |  | **Female** | **Male** |  | **Female** | **Male** |  | **Female** | **Male** |  | **Female** | **Male** |  | **Female** | **Male** |
| Argentina | 2015 |  | 31,015 | 33,770 |  | 13,981 | 13,541 |  | 48,110 | 46,904 |  | 6,241 | 6,928 |  | 2,871 | 2,373 |  | 15,542 | 13,786 |
| Australia | 2015 |  | 20,428 | 26,122 |  | 5,113 | 5,748 |  | 23,455 | 21,937 |  | 2,932 | 3,204 |  | 4,559 | 3,901 |  | 2,009 | 1,492 |
| Belgium | 2015 |  | 12,743 | 15,864 |  | 2,906 | 3,860 |  | 17,184 | 14,158 |  | 1,234 | 1,024 |  | 3,193 | 2,398 |  | 2,823 | 2,290 |
| Canada | 2013 |  | 23,246 | 27,018 |  | 3,110 | 3,969 |  | 8,905 | 17,598 |  | 1,527 | 2,562 |  | 2,129 | 2,459 |  | 747 | 1,024 |
| Chile | 2015 |  | 12,821 | 14,045 |  | 3,159 | 3,331 |  | 14,090 | 14,231 |  | 3,061 | 2,915 |  | 2,131 | 1,665 |  | 1,827 | 1,862 |
| Colombia | 2015 |  | 21,600 | 20,954 |  | 8,264 | 8,764 |  | 34,015 | 35,834 |  | 5,467 | 5,190 |  | 1,756 | 1,993 |  | 4,631 | 5,122 |
| Croatia | 2016 |  | 6,124 | 8,236 |  | 767 | 1,058 |  | 13,136 | 10,054 |  | 1,440 | 1,018 |  | 599 | 512 |  | 300 | 373 |
| Cyprus | 2016 |  | 546 | 778 |  | 182 | 238 |  | 871 | 981 |  | 146 | 204 |  | 116 | 107 |  | 38 | 39 |
| Czechia | 2016 |  | 12,421 | 15,385 |  | 1,638 | 2,344 |  | 25,354 | 22,257 |  | 2,290 | 1,937 |  | 1,724 | 1,432 |  | 1,515 | 1,671 |
| Denmark | 2015 |  | 7,430 | 8,284 |  | 2,091 | 1,957 |  | 6,215 | 6,289 |  | 734 | 900 |  | 1,237 | 1,019 |  | 947 | 866 |
| Ecuador | 2015 |  | 5,520 | 5,223 |  | 1,244 | 1,621 |  | 7,186 | 7,940 |  | 3,010 | 2,690 |  | 669 | 680 |  | 1,832 | 1,957 |
| Estonia | 2015 |  | 1,825 | 2,077 |  | 91 | 180 |  | 4,808 | 3,157 |  | 102 | 74 |  | 175 | 149 |  | 117 | 161 |
| Finland | 2015 |  | 3,583 | 4,554 |  | 303 | 573 |  | 2,121 | 5,030 |  | 115 | 187 |  | 847 | 989 |  | 36 | 52 |
| France | 2014 |  | 69,577 | 93,633 |  | 9,685 | 11,878 |  | 73,312 | 62,878 |  | 6,936 | 6,918 |  | 20,929 | 13,145 |  | 7,218 | 6,233 |
| Greece | 2015 |  | 11,814 | 18,438 |  | 3,856 | 4,192 |  | 24,272 | 22,140 |  | 1,672 | 1,798 |  | 1,508 | 1,455 |  | 3,758 | 3,071 |
| Hungary | 2016 |  | 15,283 | 18,334 |  | 2,673 | 3,314 |  | 34,927 | 27,916 |  | 1,612 | 1,329 |  | 981 | 936 |  | 624 | 642 |
| Iceland | 2016 |  | 332 | 350 |  | 65 | 54 |  | 337 | 401 |  | 15 | 29 |  | 155 | 102 |  | 40 | 30 |
| Iraq | 2008 |  | 3,835 | 4,186 |  | 797 | 1,069 |  | 20,445 | 22,685 |  | 2,668 | 2,789 |  | 657 | 851 |  | 1,060 | 1,508 |
| Ireland | 2014 |  | 4,380 | 4,840 |  | 1,119 | 1,219 |  | 4,404 | 4,448 |  | 281 | 350 |  | 763 | 657 |  | 669 | 500 |
| Israel | 2015 |  | 3,439 | 3,865 |  | 324 | 581 |  | 1,292 | 2,204 |  | 542 | 794 |  | 256 | 406 |  | 149 | 214 |
| Japan | 2015 |  | 156,230 | 225,443 |  | 32,928 | 51,478 |  | 178,744 | 160,298 |  | 14,092 | 14,973 |  | 17,068 | 13,841 |  | 57,760 | 68,179 |
| Jordan | 2012 |  | 1,155 | 1,340 |  | 115 | 246 |  | 1,902 | 2,929 |  | 550 | 652 |  | 78 | 129 |  | 113 | 175 |
| Latvia | 2015 |  | 2,898 | 3,093 |  | 97 | 254 |  | 9,561 | 6,573 |  | 380 | 222 |  | 203 | 171 |  | 154 | 294 |
| Lithuania | 2016 |  | 3,745 | 4,600 |  | 218 | 472 |  | 13,386 | 9,717 |  | 219 | 185 |  | 319 | 289 |  | 274 | 502 |
| Luxembourg | 2015 |  | 519 | 561 |  | 111 | 104 |  | 626 | 536 |  | 53 | 54 |  | 102 | 81 |  | 60 | 45 |
| Malta | 2015 |  | 427 | 510 |  | 62 | 119 |  | 691 | 643 |  | 91 | 102 |  | 50 | 49 |  | 96 | 72 |
| Mexico | 2015 |  | 42,703 | 40,891 |  | 15,971 | 18,377 |  | 78,408 | 84,387 |  | 53,647 | 52,009 |  | 5,016 | 6,109 |  | 9,506 | 12,297 |
| Netherlands | 2016 |  | 21,626 | 25,350 |  | 3,963 | 4,328 |  | 20,499 | 18,148 |  | 1,986 | 1,845 |  | 4,683 | 3,686 |  | 1,973 | 1,735 |
| Panama | 2015 |  | 1,496 | 1,735 |  | 451 | 454 |  | 2,307 | 2,861 |  | 846 | 859 |  | 299 | 275 |  | 399 | 478 |
| Paraguay | 2014 |  | 1,924 | 2,129 |  | 349 | 614 |  | 3,167 | 3,598 |  | 1,463 | 1,237 |  | 185 | 216 |  | 397 | 453 |
| Peru | 2015 |  | 10,076 | 9,275 |  | 3,658 | 4,079 |  | 8,498 | 8,868 |  | 3,143 | 3,125 |  | 166 | 239 |  | 6,799 | 7,553 |
| Philippines | 2011 |  | 26,407 | 27,044 |  | 9,347 | 18,627 |  | 77,820 | 99,538 |  | 13,751 | 14,779 |  | 2,645 | 3,405 |  | 31,769 | 41,187 |
| Poland | 2015 |  | 47,241 | 58,233 |  | 3,764 | 5,730 |  | 97,055 | 83,288 |  | 4,789 | 3,563 |  | 2,989 | 2,448 |  | 7,242 | 8,080 |
| Portugal | 2014 |  | 10,808 | 15,934 |  | 2,657 | 3,211 |  | 17,712 | 14,573 |  | 2,743 | 2,186 |  | 1,932 | 1,618 |  | 3,224 | 3,183 |
| Romania | 2016 |  | 21,096 | 30,690 |  | 2,851 | 5,059 |  | 79,443 | 69,122 |  | 2,496 | 2,345 |  | 2,226 | 1,694 |  | 2,962 | 4,653 |
| Slovakia | 2014 |  | 5,978 | 7,650 |  | 395 | 683 |  | 12,349 | 10,540 |  | 677 | 550 |  | 620 | 527 |  | 766 | 916 |
| Slovenia | 2015 |  | 1,659 | 2,503 |  | 94 | 160 |  | 909 | 1,582 |  | 81 | 129 |  | 97 | 120 |  | 47 | 85 |
| South Korea | 2015 |  | 29,852 | 48,428 |  | 4,977 | 7,749 |  | 31,342 | 28,197 |  | 7,014 | 7,248 |  | 6,414 | 4,318 |  | 8,130 | 9,124 |
| Switzerland | 2015 |  | 7,974 | 9,834 |  | 1,302 | 1,556 |  | 11,878 | 9,715 |  | 856 | 830 |  | 2,024 | 1,640 |  | 1,001 | 767 |
| Uruguay | 2015 |  | 3,739 | 4,344 |  | 1,007 | 1,231 |  | 4,976 | 4,232 |  | 489 | 502 |  | 673 | 526 |  | 668 | 561 |

Abbreviations: TB=Tuberculosis.

Table S2. ICD-10 codes for classifying the six major causes of death and the major respiratory causes of death.

| **Cause** | | **ICD-10 codes** | |
| --- | --- | --- | --- |
| **Major causes of deaths** | |  | |
| Cancer | | C00 – D48 | |
| Circulatory diseases | | I00 – J99 | |
| Chronic respiratory diseases | | J30 – J98 | |
| Respiratory infections and Tuberculosis | | A15 – A19, J00 – J22 | |
| Diabetes | | E10 – E14, N18 | |
| Neurologic diseases | | G00 – G98 | |
| **Respiratory causes of deaths** | |  | |
|  | Acute upper respiratory infections |  | J00 – J06 |
|  | Influenza |  | J09 – J11 |
|  | Pneumonia |  | J12 – J18 |
|  | Other acute lower respiratory infections |  | J20 – J22 |
|  | Other diseases of upper respiratory tract |  | J30 – J39 |
|  | Chronic lower respiratory diseases |  | J40 – J47 |
